# Immunogenicity and safety of a third dose, and immune persistence of CoronaVac vaccine in healthy adults aged 18-59 years: interim results from a double-blind, randomized, placebo-controlled phase 2 clinical trial

**DOI:** 10.1101/2021.07.23.21261026

**Authors:** Hongxing Pan, Qianhui Wu, Gang Zeng, Juan Yang, Deyu Jiang, Xiaowei Deng, Kai Chu, Wen Zheng, Fengcai Zhu, Hongjie Yu, Weidong Yin

**Author notes:** **Corresponding to** Fengcai Zhu, Jiangsu Provincial Center for Disease Control and Prevention, Nanjing 210000, China,; Hongjie Yu, School of Public Health, Fudan University, Key Laboratory of Public Health Safety, Ministry of Education, Shanghai 200032, China,; Weidong Yin, Sinovac Biotech Co., Ltd., Beijing 100085, China,. These authors contributed equally (Hongxing Pan, Qianhui Wu, Gang Zeng). These authors jointly supervised this work (Fengcai Zhu, Hongjie Yu, Weidong Yin).

## Abstract

**Background:** Large-scale vaccination is being implemented globally with CoronaVac, an inactivated vaccine against coronavirus disease 2019 (COVID-19). Immunogenicity and safety profiles of homologous two-dose schedules have been published. We report interim results of immune persistence, and the immunogenicity and safety of a third dose of CoronaVac.

**Methods:** In this ongoing, placebo-controlled, double-blind phase 2 trial in 18-to-59-year-olds, we randomly assigned subjects, 1:1:1:1, to one of four schedules to receive a third dose, 28 days or 6 months after two two-dose regimens (14-day or 28-day apart): ***schedule 1***: days 0, 14, 42; ***schedule 2***: days 0, 14, 194; ***schedule 3***: days 0, 28, 56; ***schedule 4***: days 0, 28, 208. For each schedule, participants were randomly assigned to either a medium-dose group (3 μg per 0.5 mL of aluminum hydroxide diluent per dose), a high-dose group (6 μg), or a placebo group (2:2:1). The primary outcome was geometric mean titers (GMTs) of neutralizing antibody to live SARS-CoV-2.

**Results:** Overall, 540 participants received a third dose. In the 3 μg group, neutralizing antibody titers induced by the first two doses declined after 6-8 months to below the seropositive cutoff (GMT: 4.1 [95%CI 3.3-5.2] for ***Schedule 2*** and 6.7 [95%CI 5.2-8.6] for ***Schedule 4***). When a third dose was given 6-8 months after a second dose, GMTs assessed 14 days later increased to 137.9 [95%CI 99.9-190.4] for ***Schedule 2***, and 143.1 [95%CI 110.8-184.7] for ***Schedule 4***, approximately 3-fold above ***Schedule 1*** and ***Schedule 3*** GMTs after third doses. Similar patterns were observed for the 6 μg group. The severity of solicited local and systemic adverse reactions reported within 28 days after the third dose were grade 1 to grade 2 in all vaccination cohorts. None of the fourteen serious adverse events were considered to be related to vaccination.

**Conclusions:** A third dose of CoronaVac administered 6 or more months after a second dose effectively recalled specific immune response to SARS-CoV-2, resulting in a remarkable increase in antibody levels, and indicating that a two-dose schedule generates good immune memory. Optimizing the timing of a booster dose should take into account immunogenicity, vaccine efficacy/effectiveness, local epidemic situation, infection risk, and vaccine supply. (Funded by the National Key Research and Development Program, Beijing Science and Technology Program and National Science Fund for Distinguished Young Scholars; ClinicalTrials.gov number, NCT04352608.)

## Introduction

In response to the coronavirus disease 2019 (COVID-19) pandemic, twenty COVID-19 vaccines have been approved for use^1^ and more than 3.73 billion doses have been administered as of July 20, 2021^2^. Real-world evidence has shown that vaccination effectively reduces SARS-CoV-2 transmission^3^ and decreases COVID-19 burden of disease^4-6^. A hope and expectation are that herd immunity can be achieved through mass vaccination to curb the pandemic. Amid the ongoing effort to vaccinate target populations, two critical questions are of great interest to scientists and policy-makers developing strategies to stop the pandemic: how durable is vaccine-induced immunity, and will people need a booster doseã And if so, when will people need a booster doseã

CoronaVac (Sinovac Life Sciences, Beijing, China), an inactivated vaccine against COVID-19, has been authorized for conditional use in China^7^ and is included in the World Health Organization’s (WHO) emergency use listing^8^. CoronaVac has been administered in 26 countries^1^ and is increasing the supply of COVID-19 vaccines through COVAX^9^. In China, a total of 1.46 billion doses COVID-19 vaccines have been administered as of July 19^10^, the vast majority of which are inactivated vaccines.

Evidence from real-world use of CoronaVac with a two-dose schedule in Chile demonstrated that the vaccine effectively prevents laboratory-confirmed COVID-19, and with highest effectiveness against the more severe outcomes (hospitalization, ICU admission, and death)^11^. However, persistence of CoronaVac vaccine-induced immunity is unknown, and the immunogenicity and safety of a booster dose has not been determined. We report relevant findings from an ongoing phase 2 clinical trial of CoronaVac; we compare immunogenicity and safety of a third homologous dose given at an interval of 6 months or more following the second dose to an alternative schedule with a dose-2 to dose-3 interval of 28 days.

## Methods

### Study design and participants

Our single-center, double-blind, randomized, placebo-controlled, phase 2 clinical trial of CoronaVac was initiated in Jiangsu, China on May 3, 2020, with two two-dose schedules in the original protocol. Healthy adults aged 18-59 years were eligible for enrollment. Exclusion criteria can be found in a previous publication^12^. Eligible participants were recruited and randomly allocated (1:1) to schedules with either a 14-day interval or a 28-day interval; within each schedule group, subjects were randomly allocated to either a medium-dose group (3 μg per 0.5 mL of aluminum hydroxide diluent per dose), a high-dose group (6 μg per 0.5 mL of aluminum hydroxide diluent per dose), or a placebo group (2:2:1). Interim results on safety, tolerability, and immunogenicity for these two doses have been reported^12^.

In June, 2020, the trial protocol was amended to evaluate the immunogenicity of an additional dose either 28 days or 6 months after the second dose (randomly allocated 1:1) in each of the original study groups. Accordingly, four regimens with three doses are included in this analysis: 1) the first two doses on days 0 and 14, and a third dose on day 42 (***schedule 1***); 2) the first two doses on days 0 and 14, plus a third dose 6 months after the second dose (day 194; ***schedule 2***); 3) the first two doses on days 0 and 28, and a third dose on day 56 (***schedule 3***); 4) the first two doses on days 0, and 28, plus one-dose 6 months after the second dose (day 208; ***schedule 4***). We assessed duration of immune persistence for the first two doses and evaluated immunogenicity and safety of the third doses.

Written informed consent was obtained from participants before enrolment and before administration of the third dose. The clinical trial protocol and informed consent forms were approved by the Jiangsu Ethics Committee (JSJK2020-A021-02). This trial is registered with ClinicalTrials.gov, NCT04352608. Our study was conducted in accordance with the requirements of Good Clinical Practices of China and the International Conference on Harmonization.

### Randomization and masking

Randomization codes for each vaccination schedule cohort were generated individually; the process of assignment was described previously^12^. Participants in each cohort were randomly assigned using block randomization with a block size of five, developed with SAS software (version 9.4). Concealed random grouping allocations and blinding codes were kept in signed and sealed envelopes and were blinded to investigators, participants, and laboratory staff.

### Procedures

Vaccine or placebo was given by intramuscular injection. Procedures for the first two doses have been described previously^12^. In schedule 1, routine hematological and biochemical tests were performed for the first 30 participants before dose 3 and within 3 days after dose 3. Participants would not be assigned a third dose if the severity of hematology and biochemistry indexes was grade 2 or more. Additional exclusion criteria for administration of the third dose are provided in Appendix 1.

After successful safety observations within 3 days after dose 3 in these 30 participants, the trial could proceed and the first 30 participants in ***schedule 3*** were allowed to be given their third dose (having passed haematological and biochemical criteria). After successful safety observations within 7 days after dose 3 with no abnormalities for haematological and biochemical tests, the remaining participants in ***schedule 1*** and ***schedule 3*** groups were given dose 3. For all participants in ***schedule 2*** and ***schedule 4***, a booster dose was given on the basis of interim immunogenic results obtained 6 months after the second dose. Participants would be removed from the trial if any of the following criteria were met: 1) participant request, 2) unacceptable adverse event, 3) unacceptable health status, 4) abnormal clinical manifestations judged by the investigators, 5) other reasons considered by the investigator. The trial would be suspended under the following conditions: 1) occurrence of one or more grade 4 (local and systemic) adverse reactions related to vaccination; 2) more than 15% of the participants having grade 3 or above adverse reactions, including local reactions, systemic reactions, and vital sign changes.

To evaluate immunogenicity, blood samples were collected on days 0, 28, 42, 70, and 222 from participants in the ***schedule 1*** group; on days 0, 28, 42, 194, and 208 in the ***schedule 2*** group; on days 0, 56, 84, and 236 in the ***schedule 3*** group; and on days 0, 56, 208, and 236 in the ***schedule 4*** group. The timing of each visit is shown in Appendix 2. The immunological assessment methods and related procedures were described previously^12^. Neutralizing antibodies to live SARS-CoV-2 (virus strain SARS-CoV-2/human/CHN/CN1/2020, GenBank number MT407649.1) were quantified using a micro cytopathogenic effect assay^12^.

Collection of safety information after the third dose was conducted using the same methods as for the first two doses, described previously^12^. Participants were required to record injection-site adverse events (e.g., pain, redness, swelling), or systemic adverse events (e.g., allergic reaction, cough, fever) on diary cards within 7 days after the third dose. From days 8-28 after the third dose, unsolicited adverse reactions were collected by spontaneous reporting from participants. Serious adverse events were collected until 6 months after the second dose for ***schedule 2*** and ***4*** groups, and until 6 months after three doses for ***schedule 1*** and ***3*** groups. Reported adverse events were graded according to China National Medical Products Administration guidelines^12^. Existence of causal associations between adverse events and vaccination was determined by the investigators.

### Outcomes

The primary outcome was geometric mean titers (GMTs), seropositivity, and seroconversion rate of neutralizing antibodies to live SARS-CoV-2. We defined seropositivity as a titer of 8 or higher for neutralizing antibodies to live SARS-CoV-2. Seroconversion was defined as a change of titers from seronegative at baseline to seropositive, or a four-fold increase of titers for individuals whose titers were above seropositive cutoffs.

A secondary analysis of safety endpoints included any adverse reactions within 28 days after dose 3, and serious adverse events coded by Medical Dictionary for Regulatory Activities (MedDRA) System Organ Class during the aforementioned observation period.

### Statistical analyses

We assessed immunogenic endpoints in the per-protocol population (datasets at each visit are described in Appendix 3), which included all participants who completed their assigned vaccinations. Incidence of serious adverse events was evaluated in a safety population who received at least one dose of study vaccine from the beginning of the vaccination schedule; incidence of adverse reactions was evaluated in the safety population for each dose.

We used Pearson χ^2^ test or Fisher’s exact test to analyze categorical outcomes. We calculated 95% CIs for all categorical outcomes using the Clopper-Pearson method. We calculated GMTs and corresponding 95% CIs on the basis of standard normal distribution of log-transformed antibody titers. We used t-test to compare log-transformed antibody titers between groups. Hypothesis testing was two-sided and we considered p values of less than 0.05 to be significant. We used R software (version 3.6.0) for all analyses.

The clinical trial is supervised by an independent data monitoring committee, which consists of one independent statistician, one clinician, and one epidemiologist.

## Results

### Participants

We randomly assigned 150 participants to each schedule group, and within each schedule group, 60 participants were assigned at random to a 3 μg group, 60 participants were assigned at random to a 6 μg group, and 30 participants were assigned at random to a placebo group; in all, 540 participants were eligible and allocated to receive third doses. In the ***schedule 2*** and ***schedule 4*** cohorts, 147 and 138 participants were followed up for 6 months after the second dose, and 141 and 130 participants were given third doses at 180-to-240-day intervals after the second dose for immunogenetic evaluations. In the ***schedule 1*** and ***schedule 3*** cohorts, 139 and 130 participants received third doses, and 135 and 124 participants completed blood sampling to assess immune persistence for 6 months after dose 3 (Figure 1).

**Figure 1.**
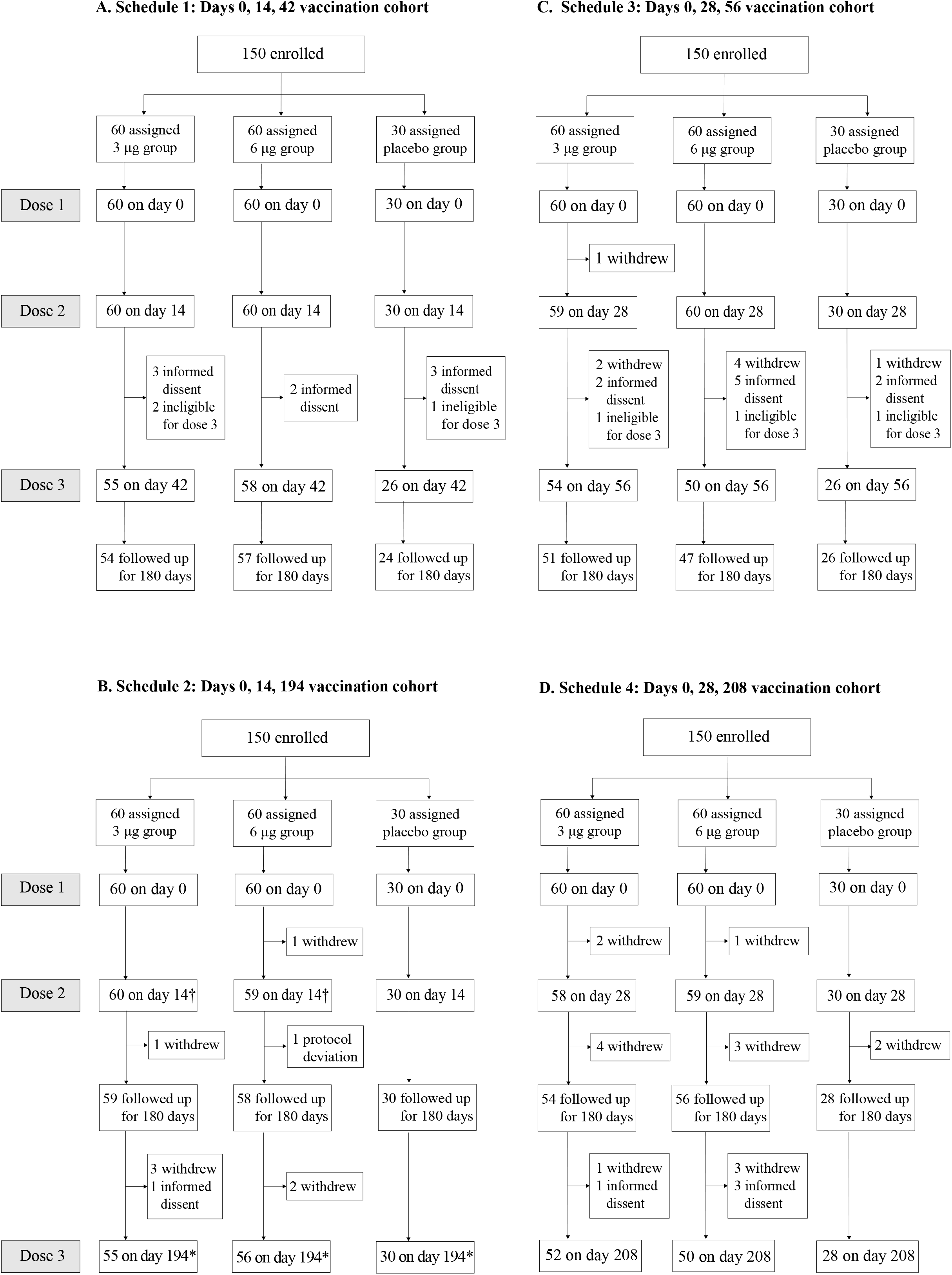
Study Profile. † One participant received the second dose on day 13, which was one day ahead of the schedule specified in the protocol. *Participants received the third dose between day 249 and day 251, exceeding the time window specified in the protocol (day 180+60).

A subject in the 6 μg group of ***schedule 2*** received a second dose of placebo by mistake and was excluded from immunogenicity analyses. There were 143 minor protocol deviations in ***schedule 2***, including two subjects given second doses at a 13-day interval after the first dose (instead of 14 days) and 141 subjects given third doses at intervals from 249-251 days after the second dose. All subjects were included in immunogenicity analyses and analyzed according to their actual situations. The mean age of participants was 44.1 years, 40.8 years, 41.2 years, 42.2 years in the four schedule groups (***schedules 1-4***). More women (53.8%) than men participated. Baseline characteristics are shown in Table 1.

**Table 1.** Baseline demographic characteristics for the safety population.

|  | 3 µg group | 6 µg group | Placebo group |
| --- | --- | --- | --- |
| **Days 0, 14, 42 vaccination cohort (*schedule 1*)** |  |  |  |
| Participants | 60 | 60 | 30 |
| Age, years | 44.9±8.9 | 43.7±8.8 | 43.5±8.3 |
| Sex (Male/Female) | 29/31 | 21/39 | 13/17 |
| **Days 0, 14, 194 vaccination cohort (*schedule 2*)** |  |  |  |
| Participants | 60 | 60 | 30 |
| Age, years | 39.0±10.7 | 41.1±9.1 | 43.6±7.0 |
| Sex (Male/Female) | 25/35 | 27/33 | 12/18 |
| **Days 0, 28, 56 vaccination cohort (*schedule 3*)** |  |  |  |
| Participants | 60 | 60 | 30 |
| Age, years | 41.0±9.1 | 40.1±9.9 | 43.9±7.4 |
| Sex (Male/Female) | 36/24 | 33/27 | 12/18 |
| **Days 0, 28, 208 vaccination cohort (*schedule 4*)** |  |  |  |
| Participants | 60 | 60 | 30 |
| Age, years | 42.0±10.2 | 41.1±10.0 | 44.7±9.5 |
| Sex (Male/Female) | 27/33 | 30/30 | 12/18 |

### Immunogenicity

At baseline, none of the participants had detectable neutralizing antibodies in the 3 μg, or 6 μg groups, regardless of schedule. In the placebo group, other than 2 of 30 (6.67%) participants in ***schedule 2***, no participants had detectable neutralizing antibodies at any of the blood-drawing visits. Given that the 3 μg formulation is the licensed formulation, we present results for the 3 μg group in the main text and provide detailed results for the 6 μg group in the Supplement.

On day 28 after the first two doses, GMTs were consistent between ***Schedule 1*** and ***Schedule 2*** with a dose spacing of 14 days (22.2 [95%CI 17.8-27.8] vs. 25.6 [95%CI 20.9-31.4]). Compared with these groups, higher GMTs were observed in both ***Schedule 3*** and ***Schedule 4*** with their longer dose spacing of 28 days (39.6 [95%CI 30.1-52.2] and 49.1 [95%CI 40.1-60.2]). However, for all two-dose schedules, regardless of dose spacing, neutralizing antibody titers declined to below the seropositive cutoff after 6 or more months (GMT: 4.1 [95%CI 3.3-5.2] for ***Schedule 2***, and 6.7 [95%CI 5.2-8.6] for ***Schedule 4***). Even though a third dose on day 28 after the first two doses slightly-to-moderately increased neutralizing antibody levels (GMT on day 28 after dose 3: 45.8 [95%CI 35.7-58.9] for ***Schedule 1***, and 49.7 [95%CI 39.9-61.9] for ***Schedule 3***), neutralizing antibodies decayed to close to the seropositive cutoff 6 months later. If a third dose was given 6 months or more after the second dose, the GMT assessed 14 days after the third dose increased to 137.9 [95%CI 99.9-190.4] for ***Schedule 2***, and 143.1 [95%CI 110.8-184.7] for ***Schedule 4***), approximately 3-fold above that of ***Schedule 1*** and ***Schedule 3*** after their third doses (Figure 2, Table S1).

**Figure 2.**
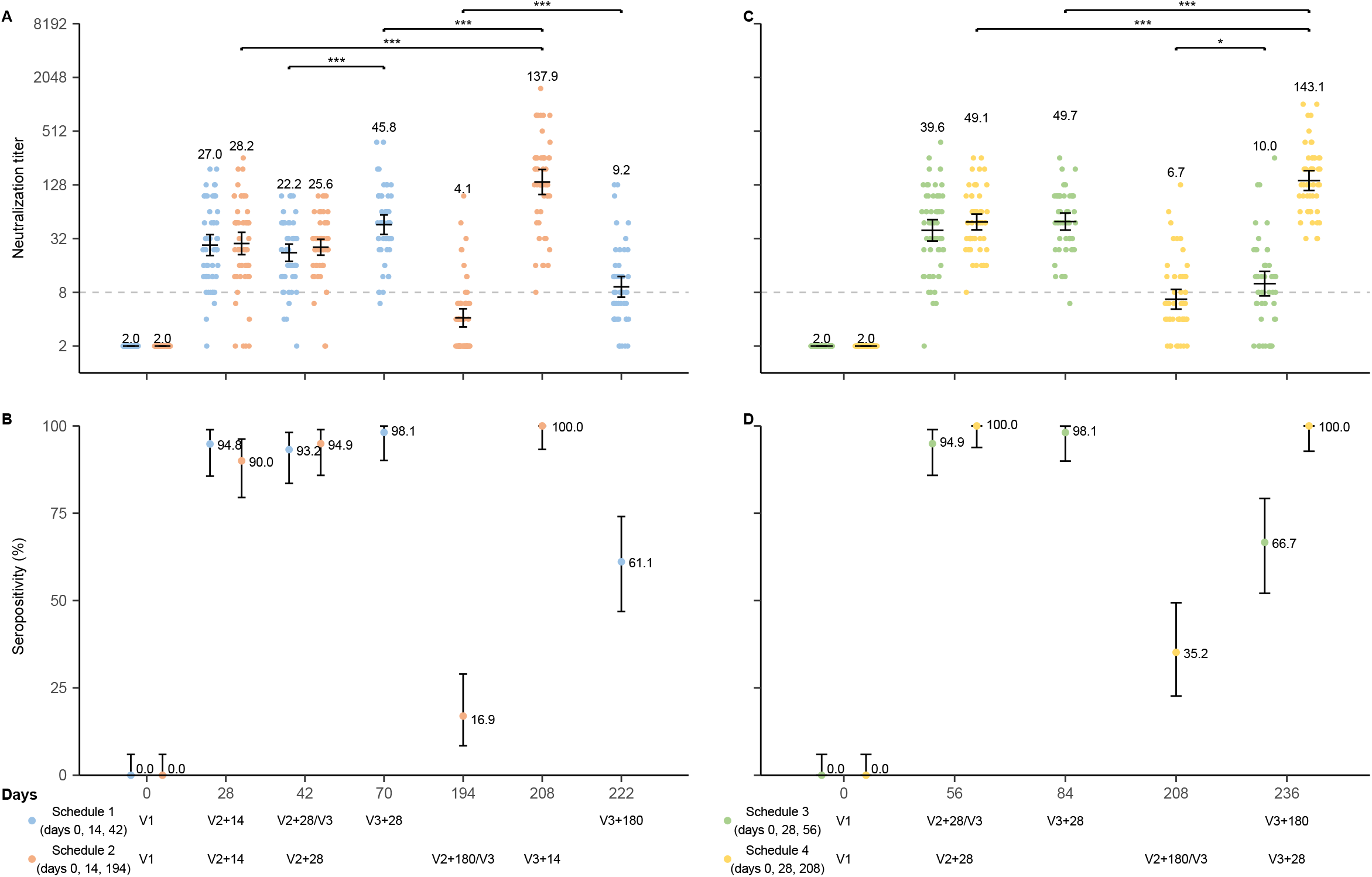
Level of neutralizing antibodies to live SARS-CoV-2 in 3 μg group: A)-B) days 0 and 14 vaccination cohort; C)-D) days 0 and 28 vaccination cohort. Note: Data are represented as reciprocal neutralizing antibody titers regarding the time after the first dose in per-protocol population. Numbers above the bars show the Geometric Mean Titer (GMT), and the error bars indicate the 95% CI. Statistical differences were assessed by t-test on log-transformed data. *p<0.05, **p<0.005, ***p<0.0005, ****p<0.0001.

Seropositivity in all four schedules was above 90.0% on day 28 after both the second dose and third dose. On day 180 after the third dose for ***schedule 1*** and ***schedule 3***, seropositivity remained above 60% while only 16.9% and 35.2% of participants in ***schedule 2*** and ***schedule 4*** were seropositive 180 days after the second dose (Figure 2, Table S1). The seroconversion rate on day 14 after the third dose for ***schedule 2*** was 100 % and on day 28 after the third dose for ***schedule 4*** was 95.9% (using neutralization antibody level before the third dose as a baseline) (Table S3).

Similar patterns were observed for the 6 μg group. Significant differences in GMT were observed between the 3 μg and 6 μg group in only a few visits for the four schedules during the study time (Table S2, Figure S1, Figure S2).

### Reactogenicity and safety

The severity of solicited local and systemic adverse reactions reported within 28 days after the third dose were grade 1 to grade 2 in all vaccination cohorts. The most common reported reaction was injection-site pain (Figure 3). Incidences of adverse reactions after the third dose were 7.91% and 3.08% for ***schedule 1*** and ***schedule 3***, lower than the overall incidence of adverse reactions within 28 days after three doses (33.33% for ***schedule 1*** and 22.00% for ***schedule 3***). In the 3 μg group, the overall incidence of adverse reactions with 28 days after the third dose was ten (18.18%) of 55 participants in ***schedule 2*** and ten (19.23%) of 52 in ***schedule 4***, which was similar with the highest incidence of adverse reactions for ***schedule 1*** (18.33% after the first dose) and ***schedule 3*** (18.33% after the first dose). Most adverse reactions were grade 1 in severity. There were no significant differences among the 3 μg, 6 μg, and placebo groups for all schedules (Table S4-S11).

**Figure 3.**
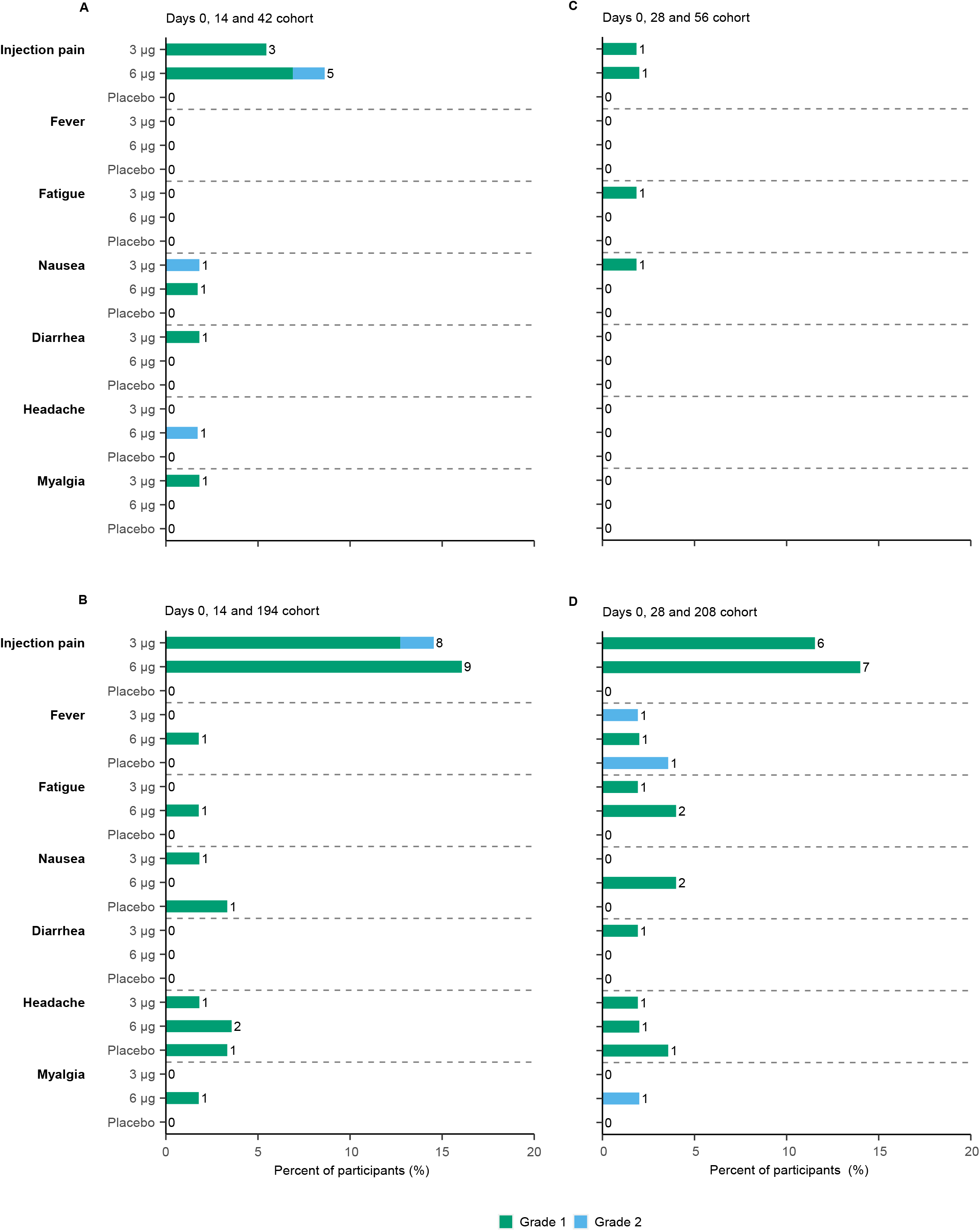
Incidence of selected adverse reactions within 28 days after the third dose.

A total of fourteen serious adverse events among nine participants were reported from the beginning of vaccination to 6 months after the second dose for ***schedule 2*** and ***4***, and to 6 months after the third dose for ***schedule 1*** and ***3*** (Table S12). None of the serious adverse events were considered by the investigators to be related with vaccination.

## Discussion

Our results demonstrated that two doses of CoronaVac (3 μg formulation) induce good immunogenicity. Although neutralizing antibody levels declined to near the positive cutoff titer of 8 after 6 months, a two-dose vaccination schedule generated good immune memory. A third dose, given at an interval of 6-8 months after the second dose, led to a strong boost in immune response, with GMTs increasing to approximately 140. Such an increase corresponds to 3-5 fold increase in neutralizing antibody titers 28 days after the second dose, indicating an anamnestic memory B cell response^13^.

SARS-CoV-2 spike-specific memory B cells are detectable in most COVID-19 patients and in all SARS-CoV-2 naïve subjects after receiving a second dose of mRNA vaccine^14,15^. The third dose of CoronaVac effectively boosts neutralizing titers and potentially provides better immuno-protection. This pattern is consistent with a recent study that reported booster immunization of AstraZeneca ChAdOx1 nCoV-19, showing higher concentrations of total IgG antibodies after a third dose^16^. We found that giving a third dose too early (28 days after the second dose) induced a much lower antibody level - only one third compared with a third dose given 6 or more months after a second dose. As shown in COVID-19 patients, memory B cells against SARS-CoV-2 spike were more abundant at 6 months than at 1 month after symptom onset^14^.

For the Moderna mRNA-1273 vaccine, antibody declined slightly as expected, but remained high in all ages on days 90 and 180 after the second dose, with antibody detected among all participants^17,18^. For Pfizer BNT162b2 and AstraZeneca ChAdOx1 nCoV-19 vaccines, antibody levels declined by 55% and 84% between 21–41 days and 70 days or more after a second dose^19^. However, it is difficult to directly compare these estimates with our findings for CoronaVac due to heterogeneity of neutralization assays. Even with neutralization assays that use the same live virus, given the lack of standardized laboratory methods for SARS-CoV-2 neutralization and experimental procedures, including virus titration, serum dilution, virus-serum neutralization, readout, and reporting method (e.g., NT50, NT100), results vary greatly by laboratory^20^.

During the first three months of Chile’s mass vaccination campaign, a two-dose CoronaVac schedule showed good effectiveness against COVID-19: 65.9% for symptomatic infection, 87.5% for hospitalization, 90.3% for ICU admission, and 86.3% for death among a population aged 16 years or older^11^. Our study demonstrated that the neutralizing antibody level decayed to around the positive cutoff of 8 by 6-8 months after the second dose. Vaccine effectiveness in this case is unclear, since the protection threshold of titers against COVID-19 remains unknown, and both T cell immunity and B cell memory against SARS-CoV-2 elicited by inactivated vaccines may contribute to protection^13,21^. In addition, establishment of SARS-CoV-2 spike-specific immune memory, other than inducing durable antibody, may be important for a successful COVID-19 vaccine^12^. In epidemic areas with SARS-CoV-2 circulation, natural infection after two-dose vaccination may play the role of a booster dose^22^. Accordingly, although a third dose of CoronaVac would be essential, the timing of a booster dose must account for the local epidemic situation, risk of infection, vaccine supply, and other relevant factors. In the short-to-medium term, ensuring more people complete the current two-dose schedule of CoronaVac should be the priority.

CoronaVac is approved for use in a two-dose schedule with an interval of 14-28 days between doses. In our study, dose spacing impacted immunogenicity. A longer interval between the first and second doses triggered a stronger immune response (28 days vs. 14 days: about 2-fold GMT on day 28 after the second dose, and 2-fold seropositivity at 6 months after the second dose), as has been seen with other COVID-19 vaccines, such as ChAdOx1-S, and BNT162b2^23,24^.

The incidence of adverse reactions after the third dose was lower than the highest incidence of adverse reactions during the observation period in a previous study^12^. That study found a 24.2% incidence after the first dose in the days 0 and 14 schedule, indicating that a third dose was well-tolerated. Although no major safety concern was identified during our trial, adverse reactions are important to monitor in large-scale clinical trials and during post-marketing periods.

Our study had several limitations. First, T cell responses were not assessed in the phase 2 trial. Second, we only reported immune response data for healthy adults, and did not include individuals who are more susceptible, have higher risk of severe outcomes, but have lower neutralizing antibody titers after two-dose vaccination (e.g., older individuals [aged≥60 years] or with comorbidities)^25,26^. Data on immune persistence needs further study, particularly immune persistence of a third dose given 6 months or more after the second dose. Third, although there were no tolerability concerns reported from those receiving a third dose, the sample size in our study is not sufficiently large to assess rare vaccine side effects. Fourth, we did not perform neutralization test in vitro against variants to determine the neutralization ability of the vaccine to emerging variants of concern.

In summary, our study found that a two-dose vaccination schedule of CoronaVac (3 μg formulation) generated good immune memory. Although the neutralizing antibody titer dropped to low levels 6 months after the second dose, a third dose was highly effective at recalling a SARS-CoV-2-specific immune response, leading to a significant rebound in antibody levels. Determining the timing of a booster dose must take into account many factors, including immunogenicity, vaccine efficacy/effectiveness, the epidemic situation, risk of infection, and vaccine supply.

## Data Availability

The results of this study are preliminary and the study is ongoing. Access to patient-level data with qualified external researchers may be available upon request once the trial is complete.

## Acknowledgements

We thanked Dr. Lawrence Everett Rodewald from Chinese Center for Disease Control and Prevention for his kind comments on this paper.

## References

1. COVID-19 Vaccine Market Dashboard. https://www.unicef.org/supply/covid-19-vaccine-market-dashboard Accessed 21 July 2021.

2. Our world in Data. https://ourworldindata.org/covid-vaccinations Accessed 22 July 2021.

3. Harris RJ, Hall JA, Zaidi A, Andrews NJ, Dunbar JK, Dabrera G. Effect of Vaccination on Household Transmission of SARS-CoV-2 in England. The New England journal of medicine 2021.

4. Dagan N, Barda N, Kepten E, et al. BNT162b2 mRNA Covid-19 Vaccine in a Nationwide Mass Vaccination Setting. New England Journal of Medicine 2021;384:1412–23.

5. Haas EJ, Angulo FJ, McLaughlin JM, et al. Impact and effectiveness of mRNA BNT162b2 vaccine against SARS-CoV-2 infections and COVID-19 cases, hospitalisations, and deaths following a nationwide vaccination campaign in Israel: an observational study using national surveillance data. Lancet (London, England) 2021;397:1819–29.

6. Vasileiou E, Simpson CR, Shi T, et al. Interim findings from first-dose mass COVID-19 vaccination roll-out and COVID-19 hospital admissions in Scotland: a national prospective cohort study. Lancet (London, England) 2021;397:1646–57.

7. China Food and Drug Administration. Conditional use approval for CoronaVac. https://www.nmpagovcn/yaopin/ypjgdt/20210206154636109html.

8. Kremsner P, Mann P, Bosch J, et al. Phase 1 Assessment of the Safety and Immunogenicity of an mRNA-Lipid Nanoparticle Vaccine Candidate Against SARS-CoV-2 in Human Volunteers. medRxiv 2020:2020.11.09.20228551.

9. COVAX Global Supply Forecast. July 12, 2021. https://www.gavi.org/sites/default/files/covid/covax/COVAX-Supply-Forecast.pdf Accessed 21 July 2021.

10. COVID-19 vaccination. National Health Commission of the People’s Republic of China. http://www.gov.cn/xinwen/2021-07/20/content_5626149.htm Accessed July 21, 2021.

11. Jara A, Undurraga EA, González C, et al. Effectiveness of an Inactivated SARS-CoV-2 Vaccine in Chile. New England Journal of Medicine 2021.

12. Zhang Y, Zeng G, Pan H, et al. Safety, tolerability, and immunogenicity of an inactivated SARS-CoV-2 vaccine in healthy adults aged 18–59 years: a randomised, double-blind, placebo-controlled, phase 1/2 clinical trial. The Lancet Infectious Diseases 2021;21:181–92.

13. Quast I, Tarlinton D. B cell memory: understanding COVID-19. Immunity 2021;54:205–10.

14. Dan JM, Mateus J, Kato Y, et al. Immunological memory to SARS-CoV-2 assessed for up to 8 months after infection. Science 2021;371.

15. Goel RR, Apostolidis SA, Painter MM, et al. Distinct antibody and memory B cell responses in SARS-CoV-2 naïve and recovered individuals following mRNA vaccination. Sci Immunol 2021;6.

16. Flaxman A, Marchevsky N, Jenkin D, Aboagye J, Aley PK. Tolerability and immunogenicity after a late second dose or a third dose of ChAdOx1 nCoV-19 (AZD1222). ssrn 2021.

17. Doria-Rose N, Suthar MS, Makowski M, et al. Antibody Persistence through 6 Months after the Second Dose of mRNA-1273 Vaccine for Covid-19. New England Journal of Medicine 2021.

18. Widge AT, Rouphael NG, Jackson LA, et al. Durability of responses after SARS-CoV-2 mRNA-1273 vaccination. New England Journal of Medicine 2021;384.

19. Shrotri M, Navaratnam AMD, Nguyen V, et al. Spike-antibody waning after second dose of BNT162b2 or ChAdOx1. Lancet (London, England) 2021.

20. Chen X, Chen Z, Azman AS, et al. Comprehensive mapping of neutralizing antibodies against SARS-CoV-2 variants induced by natural infection or vaccination. medRxiv : the preprint server for health sciences 2021.

21. Deng Y, Li Y, Yang R, Tan W. SARS-CoV-2-specific T cell immunity to structural proteins in inactivated COVID-19 vaccine recipients. Cell Mol Immunol 2021.

22. Chen H, Mao Y, Duan Z, et al. Three Cases of COVID-19 Variant Delta With and Without Vaccination — Chengdu City, Sichuan Province, April–May, 2021. China CDC Weekly 2021;3:544–6.

23. Voysey M, Clemens SAC, Madhi SA, et al. Single-dose administration and the influence of the timing of the booster dose on immunogenicity and efficacy of ChAdOx1 nCoV-19 (AZD1222) vaccine: a pooled analysis of four randomised trials. Lancet (London, England) 2021;397:881–91.

24. Parry H, Bruton R, Stephens C, et al. Extended interval BNT162b2 vaccination enhances peak antibody generation in older people. medRxiv 2021.

25. Monin L, Laing AG, Muñoz-Ruiz M, et al. Safety and immunogenicity of one versus two doses of the COVID-19 vaccine BNT162b2 for patients with cancer: interim analysis of a prospective observational study. Lancet Oncol 2021;22:765–78.

26. Maneikis K, Šablauskas K, Ringelevičiūtė U, et al. Immunogenicity of the BNT162b2 COVID-19 mRNA vaccine and early clinical outcomes in patients with haematological malignancies in Lithuania: a national prospective cohort study. Lancet Haematol 2021.

