## Supplementary for "Immunogenicity and safety of a third dose, and immune persistence of CoronaVac vaccine in healthy adults aged 18-59 years: interim results from a double-blind, randomized, placebo-controlled phase 2 clinical trial"

Supplemental Methods

Appendix 1. Exclusion criteria for the administration of the second and third dose

- **Continuing vaccination is prohibited, and other research steps may continue according to researchers’ judgement:**

(1) The same kind of vaccine other than the experimental vaccine was used during the study period;

(2) Any serious adverse reactions with causality to the test article inoculated;

(3) Severe anaphylaxis or hypersensitivity after vaccination (including urticaria / rash within 30 minutes after vaccination);

(4) Any confirmed or suspected autoimmune or immunodeficiency disease, including human immunodeficiency virus (HIV) infection;

- **Vaccination can be postponed within the time window specified in the program:**

(5) Acute or new chronic diseases occurred after vaccination;

(6) Other reactions (including severe pain, severe swelling, severe activity limitation, persistent high fever, severe headache or other systemic or local reactions) were judged by the researcher;

- **Vaccination can be postponed within the time window specified in the protocol:**

(7) At the time of vaccination, the participant was suffering from acute disease (acute disease refers to moderate or severe disease with or without fever);

(8) The axillary temperature was higher than 37.0℃;

(9) Participants were vaccinated with subunit vaccine or inactivated vaccine within 7 days and attenuated vaccine within 14 days;

(10) According to the judgment of the researcher, the participant had any other factors not suitable for vaccination.

Appendix 2: Trial profile

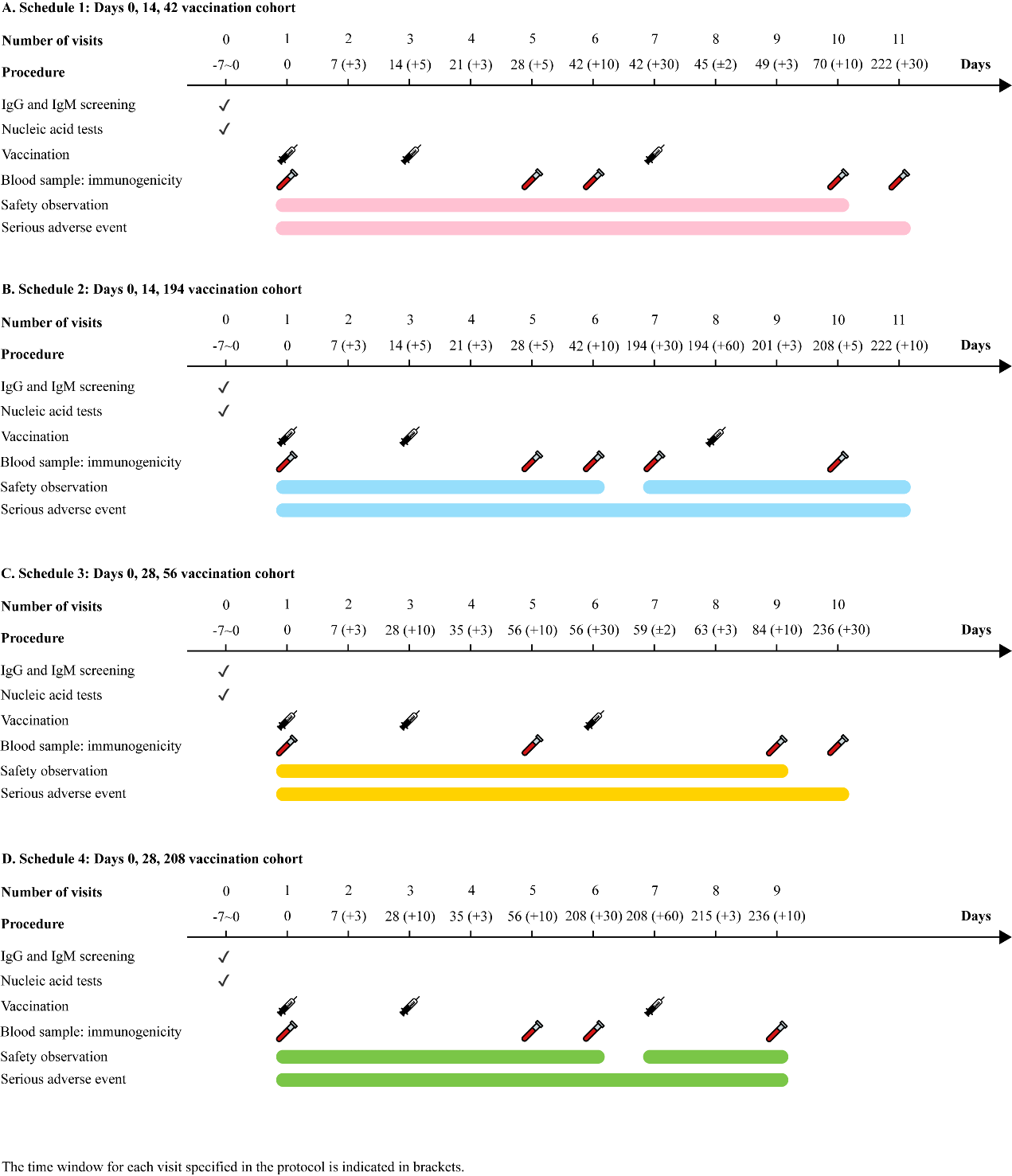

Appendix 3: Descriptions of data sets

**Full Analysis Set (FAS)**

FAS included subjects who received at least one dose of interventional product, completed at least one blood sampling before and after vaccination for immunogenicity evaluation during the whole vaccination schedule following the principle of intention to treatment (ITT). Subjects with wrong vaccination were evaluated for immunogenicity in original randomized groups according to ITT principle.

**Per Protocol Set (PPS)**

PPS was a subset of FAS, including all subjects who met enrolment criteria and received two doses of vaccines within the time window according to the protocol requirements. Blood samples of subjects were collected before the first dose and at 14 days after the second dose, and evaluated for immunogenicity effectively. Exclusion criteria for PPS was as follows:

- subjects violate protocol;
- subjects injected with wrong interventional product;
- the injection of second dose or the blood sampling at 14 days after the second dose beyond the window;
- subjects using vaccines or drugs prohibited in protocol, including
  - other research or unregistered products (drugs or vaccines) in addition of investigational product,
  - long-term use (more than 14 days) of immunosuppressants or other immunomodulatory drugs (inhaled or topical steroids are allowed),
  - immunoglobulins and / or blood preparations.
- subjects with newly diagnosed autoimmune diseases, including human immunodeficiency virus (HIV) infection;
- other factors affecting the immunogenicity evaluation of the vaccine at 14 days after the second dose.

**Per Protocol Set 2B (PPS-2B)**

PPS-2B was a subset of FAS, including all subjects who met enrolment criteria and received two doses of vaccines within the time window according to the protocol requirements. Blood samples of subjects were collected before the first dose and at 28 days after the second dose, and evaluated for immunogenicity effectively. Exclusion criteria for PPS-2B was as follows:

- subjects violate protocol;
- subjects injected with wrong interventional product;
- the injection of second dose or the blood sampling at 28 days after the second dose beyond the window;
- subjects using vaccines or drugs prohibited in protocol, including
  - other research or unregistered products (drugs or vaccines) in addition of investigational product,
  - long-term use (more than 14 days) of immunosuppressants or other immunomodulatory drugs (inhaled or topical steroids are allowed),
  - immunoglobulins and / or blood preparations.
- subjects with newly diagnosed autoimmune diseases, including human immunodeficiency virus (HIV) infection;
- other factors affecting the immunogenicity evaluation of the vaccine at 28 days after the second dose.

**Per Protocol Set 3 (PPS3)**

PPS3 was a subset of FAS, including all subjects who met enrolment criteria and received three doses of vaccines within the time window according to the protocol requirements. Blood samples of subjects were collected after the third dose and evaluated for immunogenicity effectively. Exclusion criteria for PPS3 was as follows:

- subjects violate protocol;
- subjects injected with wrong interventional product;
- the injection of second and third dose or the blood sampling at 28 days after the third dose beyond the window;
- subjects using vaccines or drugs prohibited in protocol, including
  - other research or unregistered products (drugs or vaccines) in addition of investigational product,
  - long-term use (more than 14 days) of immunosuppressants or other immunomodulatory drugs (inhaled or topical steroids are allowed),
  - immunoglobulins and / or blood preparations.
- subjects with newly diagnosed autoimmune diseases, including human immunodeficiency virus (HIV) infection;
- other factors affecting the immunogenicity evaluation of the vaccine at 28 days after the third dose.

**Immune Persistence Set (IPS-6)**

IPS-6 included subjects who completed the full course immunization and completed blood sampling for effective immunogenicity evaluation at 6 months after the full course of vaccination.

**Immune Persistence Set 6A (IPS-6A)**

IPS-6A included subjects who completed the two-dose schedule (a 14-day interval or a 28-day interval) primary immunization and completed blood sampling for effective immunogenicity evaluation at 6 months after the two-dose vaccination.

**Immune Persistence Set 6B (IPS-6B)**

IPS-6B included subjects who completed the three-dose schedule (day 0, 14, 42 or day 0, 28, 56) immunization and completed blood sampling for effective immunogenicity evaluation at 6 months after the three-dose vaccination.

**Full Analysis Set for Booster (bFAS)**

bFAS included subjects who received the booster of interventional product, completed at least one blood sampling before and after booster vaccination for immunogenicity evaluation following the principle of intention to treatment (ITT). Subjects with wrong vaccination were evaluated for immunogenicity in original randomized groups according to ITT principle.

**Per Protocol Set for Booster (bPPS)**

bPPS was a subset of bFAS, including all subjects who met enrolment criteria and received two-dose primary immunization and the booster after 6 months within the time window according to the protocol requirements. Blood samples of subjects were collected before the booster and at 14 days (days 0, 14 and 194 vaccination cohort) or 28 days (days 0, 28 and 208 vaccination cohort) after the booster dose, and evaluated for immunogenicity effectively. Exclusion criteria for bPPS was as follows:

- subjects violate protocol;
- subjects injected with wrong interventional product;
- the injection of booster dose or the blood sampling at 14 or 28 days after the booster dose beyond the window;
- subjects using vaccines or drugs prohibited in protocol, including
  - other research or unregistered products (drugs or vaccines) in addition of investigational product,
  - long-term use (more than 14 days) of immunosuppressants or other immunomodulatory drugs (inhaled or topical steroids are allowed),
  - immunoglobulins and / or blood preparations.
- subjects with newly diagnosed autoimmune diseases, including human immunodeficiency virus (HIV) infection;
- other factors affecting the immunogenicity evaluation of the vaccine at 14 or 28 days after the booster dose.

**Safety Set (SS)**

SS included subjects who received at least one dose of interventional product. Subjects with wrong vaccination were evaluated for safety in original randomized groups according to ASaT (All Subjects are Treated) principle. The total safety set is divided into safety set 1 (SS1), safety set 2 (SS2), safety set 3 (SS3) and safety set for booster (bSS) in each vaccination cohort. The safety analysis of each dose was based on the actual number of vaccinees received each dose. SS1 included subjects received the first dose; SS2 included subjects received the second dose; SS3 included subjects received the third dose who assigned with three-dose schedule (days 0, 14, 42 and days 0, 28, 56) vaccination in phase 2; bSS included subjects received the booster dose who assigned with booster schedule (days 0, 14, 194 and days 0, 28, 208) vaccination.

Supplemental Results of immunogenicity

Table S1. Level of neutralizing antibodies to live SARS-CoV-2 in 3 μg group (per-protocol analysis)

| **Days from**  **vaccination** | **Number of participants** | **Dose** | **GMT**  **(95%CI)** | **Seropositivity**  **(95%CI)** |  | **Days from**  **vaccination** | **Number of participants** | **Dose** | **GMT**  **(95%CI)** | **Seropositivity**  **(95%CI)** |  | **P value*** |
| --- | --- | --- | --- | --- | --- | --- | --- | --- | --- | --- | --- | --- |
| ***Schedule 1 (Days 0, 14, 42 vaccination cohort)*** | | | | |  | ***Schedule 3 (Days 0, 28, 56 vaccination cohort)*** | | | | |  |  |
| 0 | 60 (Baseline) | V1 | 2.0  (2.0, 2.0) | 0.0  (0.0, 6.0) |  | 0 | 60 (Baseline) | V1 | 2.0  (2.0, 2.0) | 0.0  (0.0, 6.0) |  | ― |
| 28 | 58 (PPS) | V2+14 | 27.0  (20.6, 35.4) | 94.8  (85.6, 98.9) |  | ― | ― | ― | ― | ― |  | ― |
| 42 | 59 (PPS-2B) | V2+28/V3 | 22.2  (17.8, 27.8) | 93.2  (83.5, 98.1) |  | 56 | 59 (PPS-2B) | V2+28/V3 | 39.6  (30.1, 52.2) | 94.9  (85.9-98.9) |  | 0.001 |
| 70 | 54 (PPS3) | V3+28 | 45.8  (35.7, 58.9) | 98.1  (90.1, 100.0) |  | 84 | 53 (PPS3) | V3+28 | 49.7  (39.9, 61.9) | 98.1  (89.9, 100.0) |  | 0.627 |
| 222 | 54 (IPS-6B) | V3+180 | 9.2  (7.1, 12.0) | 61.1  (46.9, 74.1) |  | 236 | 51 (IPS-6B) | V3+180 | 10.0  (7.3, 13.7) | 66.7  (52.1-79.2) |  | 0.691 |
| ***Schedule 2 (Days 0, 14, 194 vaccination cohort)*** | | | | |  | ***Schedule 4 (Days 0, 28, 208 vaccination cohort)*** | | | | |  |  |
| 0 | 60 (Baseline) | V1 | 2.0  (2.0, 2.0) | 0.0  (0.0, 6.0) |  | 0 | 60 (Baseline) | V1 | 2.0  (2.0, 2.0) | 0.0  (0.0, 6.0) |  | ― |
| 28 | 60 (PPS) | V2+14 | 28.2  (21.1, 37.6) | 90.0  (79.5, 96.2) |  | ― | ― | ― | ― | ― |  | ― |
| 42 | 59 (PPS-2B) | V2+28 | 25.6  (20.9, 31.4) | 94.9  (85.9, 98.9) |  | 56 | 58 (PPS-2B) | V2+28 | 49.1  (40.1, 60.2) | 100.0  (93.8, 100.0) |  | <0.001 |
| 194 | 59 (IPS-6A) | V2+180/V3 | 4.1  (3.3, 5.2) | 16.9  (8.4, 29.0) |  | 208 | 54 (IPS-6A) | V2+180/V3 | 6.7  (5.2, 8.6) | 35.2  (22.7, 49.4) |  | 0.006 |
| 208 | 53 (bPPS) | V3+14 | 137.9  (99.9, 190.4) | 100.0  (93.3, 100.0) |  | 236 | 49 (bPPS) | V3+28 | 143.1  (110.8, 184.7) | 100.0  (92.7, 100.0) |  | 0.858 |

*p values are for comparisons of neutralization geometric titers at the same interval after vaccination between ***schedule*** ***1*** and ***schedule*** ***3***, ***schedule*** ***2*** and ***schedule*** ***4***, respectively.

Table S2. Level of neutralizing antibodies to live SARS-CoV-2 in 6 μg group (per-protocol analysis)

| **Days from**  **vaccination** | **Number of participants** | **Dose** | **GMT**  **(95%CI)** | **Seropositivity**  **(95%CI)** |  | **Days**  **from vaccination** | **Number of participants** | **Dose** | **GMT**  **(95%CI)** | **Seropositivity**  **(95%CI)** |  | **P value*** |
| --- | --- | --- | --- | --- | --- | --- | --- | --- | --- | --- | --- | --- |
| ***Schedule 1 (Days 0, 14, 42 vaccination cohort)*** | | | | |  | ***Schedule 3 (Days 0, 28, 56 vaccination cohort)*** | | | | |  |  |
| 0 | 60 (Baseline) | V1 | 2.0  (2.0, 2.0) | 0.0  (0.0, 6.0) |  | 0 | 60 (Baseline) | V1 | 2.0  (2.0, 2.0) | 0.0  (0.0, 6.0) |  | ― |
| 28 | 60 (PPS) | V2+14 | 40.8  (31.7, 52.4) | 98.3  (91.1, 100.0) |  | ― | ― | ― | ― | ― |  | ― |
| 42 | 60 (PPS-2B) | V2+28 | 29.1  (23.7, 35.8) | 98.3  (91.1, 100.0) |  | 56 | 60 (PPS-2B) | V2+28 | 58.4  (46.9, 72.7) | 100.0  (94.0, 100.0) |  | <0.001 |
| 70 | 58 (PPS3) | V2+180/V3 | 74.2  (59.0, 93.3) | 98.3  (90.8, 100.0) |  | 84 | 48 (PPS3) | V2+180/V3 | 51.9  (41.3, 65.3) | 100.0  (92.6, 100.0) |  | 0.192 |
| 222 | 57 (IPS-6A) | V3+14 | 13.6  (10.5, 17.7) | 68.4  (54.8, 80.1) |  | 236 | 47 (IPS-6A) | V3+28 | 10.2  (7.1, 14.6) | 51.1  (36.1, 65.9) |  | 0.029 |
| ***Schedule 2 (Days 0, 14, 194 vaccination cohort)*** | | | | |  | ***Schedule 4 (Days 0, 28, 208 vaccination cohort)*** | | | | |  |  |
| 0 | 60 (Baseline) | V1 | 2.0  (2.0, 2.0) | 0.0  (0.0, 6.0) |  | 0 | 60 (Baseline) | V1 | 2.0  (2.0, 2.0) | 0.0  (0.0, 6.0) |  | ― |
| 28 | 59 (PPS) | V2+14 | 29.2  (21.8, 39.0) | 98.3  (90.9, 100.0) |  | ― | ― | ― | 175.1  (138.8, 221.0) | 56.1  (42.4, 69.3) |  | ― |
| 42 | 58 (PPS-2B) | V2+28/V3 | 31.1  (25.4, 38.0) | 100.0  (93.8, 100.0) |  | 56 | 58 (PPS-2B) | V2+28/V3 | 73.6  (60.2, 90.0) | 100.0  (93.8, 100.0) |  | <0.001 |
| 194 | 58 (IPS-6A) | V3+28 | 4.8  (3.8, 6.1) | 24.1  (13.9, 37.2) |  | 208 | 56 (IPS-6A) | V3+28 | 7.1  (5.6, 8.9) | 46.4  (33.0, 60.3) |  | 0.022 |
| 208 | 55 (bPPS) | V3+180 | 175.1  (138.8, 221.0) | 100.0  (93.5, 100.0) |  | 236 | 48 (bPPS) | V3+180 | 215.7  (162.6, 286.2) | 100.0  (92.6, 100.0) |  | 0.256 |

*p values are for comparisons of neutralization geometric titers at the same interval after vaccination between ***schedule*** ***1*** and ***schedule*** ***3***, ***schedule*** ***2*** and ***schedule*** ***4***, respectively.

Table S3. Seroconversion rates of neutralizing antibodies to live SARS-CoV-2

|  | **3 μg group** | **6 μg group** | **Placebo group** | **p value*** | **p value†** |
| --- | --- | --- | --- | --- | --- |
| **Schedule 1: Days 0, 14, 42 vaccination cohort** | | | | |  |
| Day 14 after 2nd dose (day 28) | 55/58 (94.8) (85.6, 98.9) | 59/60 (98.3) (91.1, 100.0) | 0/30 (0.0) (0.0, 11.6) | <0.0001 | 0.3601 |
| Day 28 after 2nd dose (day 42) | 55/59 (93.2) (83.5, 98.1) | 59/60 (98.3) (91.1, 100.0) | 0/30 (0.0) (0.0, 11.6) | <0.0001 | 0.2068 |
| Day 28 after 3rd dose (day 70) | 53/54 (98.1) (90.1, 100.0) | 57/58 (98.3) (90.8, 100.0) | 0/26 (0.0) (0.0,13.2) | <0.0001 | 1.0000 |
| **Schedule 2: Days 0, 14, 194 vaccination cohort** | | | | |  |
| Day 14 after 2nd dose (day 28) | 54/60 (90.0) (79.5, 96.2) | 58/59 (98.3) (90.9, 100.0) | 2/30 (6.7) (0.8, 22.1) | <0.0001 | 0.1140 |
| Day 28 after 2nd dose (day 42) | 56/59 (94.9) (85.9, 98.9) | 58/58 (100.0) (90.9, 100.0) | 0/30 (0.0) (0.0, 11.6) | <0.0001 | 0.2436 |
| Day 14 after 3rd dose‡ (day 208) | 52/53 (98.1)  (89.9, 100.0) | 55/55 (100.0) (93.5, 100.0) | 0/30 (0.0) (0.0, 11.6) | <0.0001 | 0.4907 |
| **Schedule 3: Days 0, 28, 56 vaccination cohort** | | | | |  |
| Day 28 after 2nd dose (day 56) | 56/59 (94.9) (85.6, 98.9) | 60/60 (100.00) (94.0, 100.0) | 0/30 (0.0) (0.0, 11.6) | <0.0001 | 0.1187 |
| Day 28 after 3rd dose (day 84) | 52/53 (98.1) (89.9, 100.0) | 48/48 (100.0) (92.6, 100.0) | 0/25 (0.0) (0.0,13.7) | <0.0001 | 1.0000 |
| **Schedule 4: Days 0, 28, 208 vaccination cohort** | | | | |  |
| Day 28 after 2nd dose (day 56) | 58/58 (100.0) (93.8, 100.0) | 58/58 (100.0) (93.8, 100.0) | 0/29 (0.00) (0.0, 11.9) | <0.0001 | 1.0000 |
| Day 28 after 3rd dose‡ (day 236) | 47/49 (95.92) (86.0, 99.5) | 46/48 (95.8) (85.8, 99.5) | 0/27 (0.0) (0.00,12.8) | <0.0001 | 1.0000 |

Data are n/N (%; 95% CI). *p values are for comparisons among three groups. †p values are for comparisons between the 3 μg and 6 μg groups. ‡Seroconversion rate was calculated based on the neutralization titer assessed before the third dose.

Figure S1. Level of neutralizing antibodies to live SARS-CoV-2 in 6 μg group: A)-B) days 0 and 14 vaccination cohort; C)-D) days 0 and 28 vaccination cohort

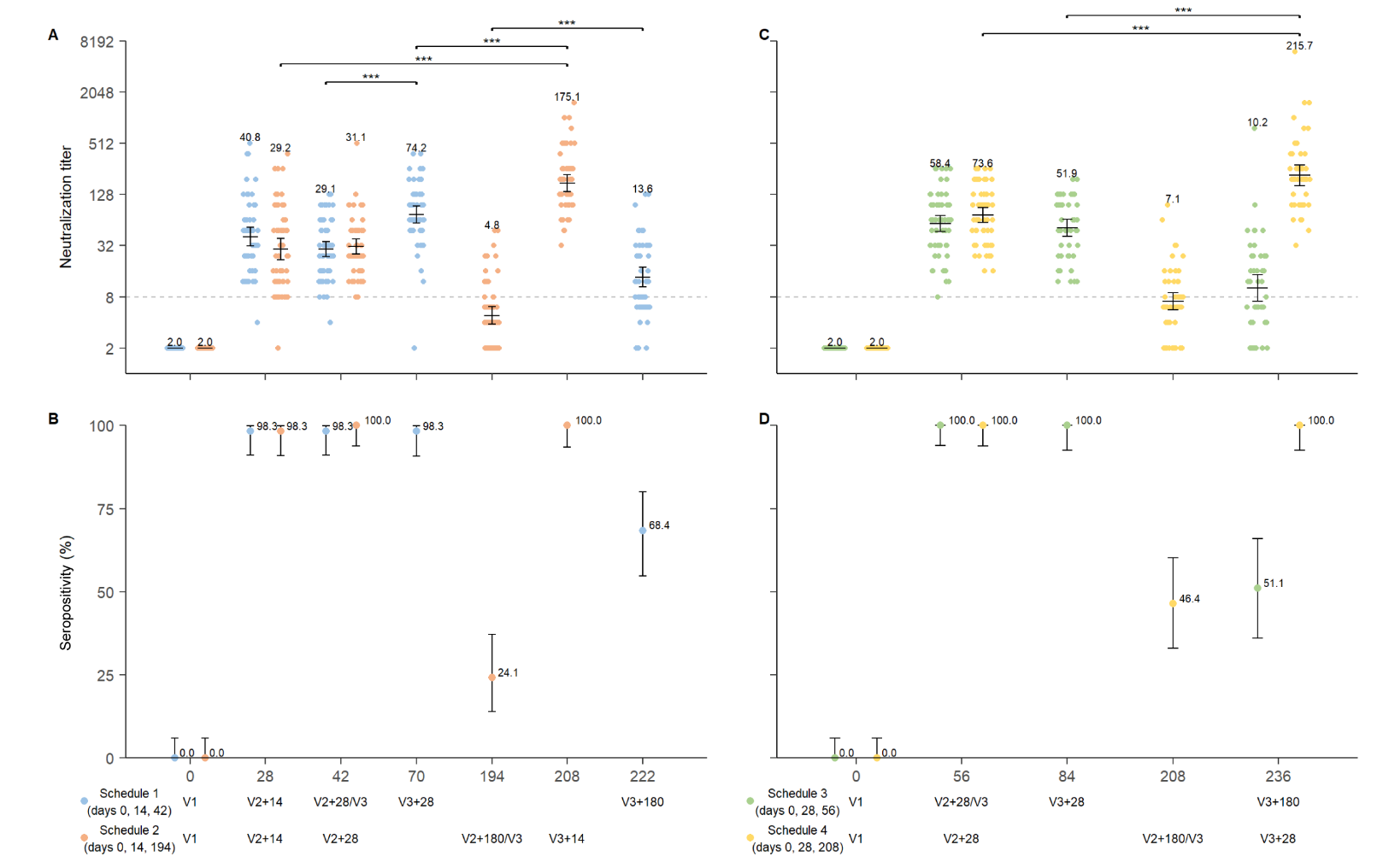

Note: Data is represented as the reciprocal neutralizing antibody titer regarding the time after the first dose in per-protocol population. Numbers above the bars show the Geometric Mean Titer (GMT), and the error bars indicate the 95% CI. Statistical differences were assessed by t-test on log-transformed data. *p<0.05, **p<0.005, ***p<0.0005, ****p<0.0001.

Figure S2. Antibody titres of neutralising antibodies to live SARS-CoV-2 and seropositivity for four schedules

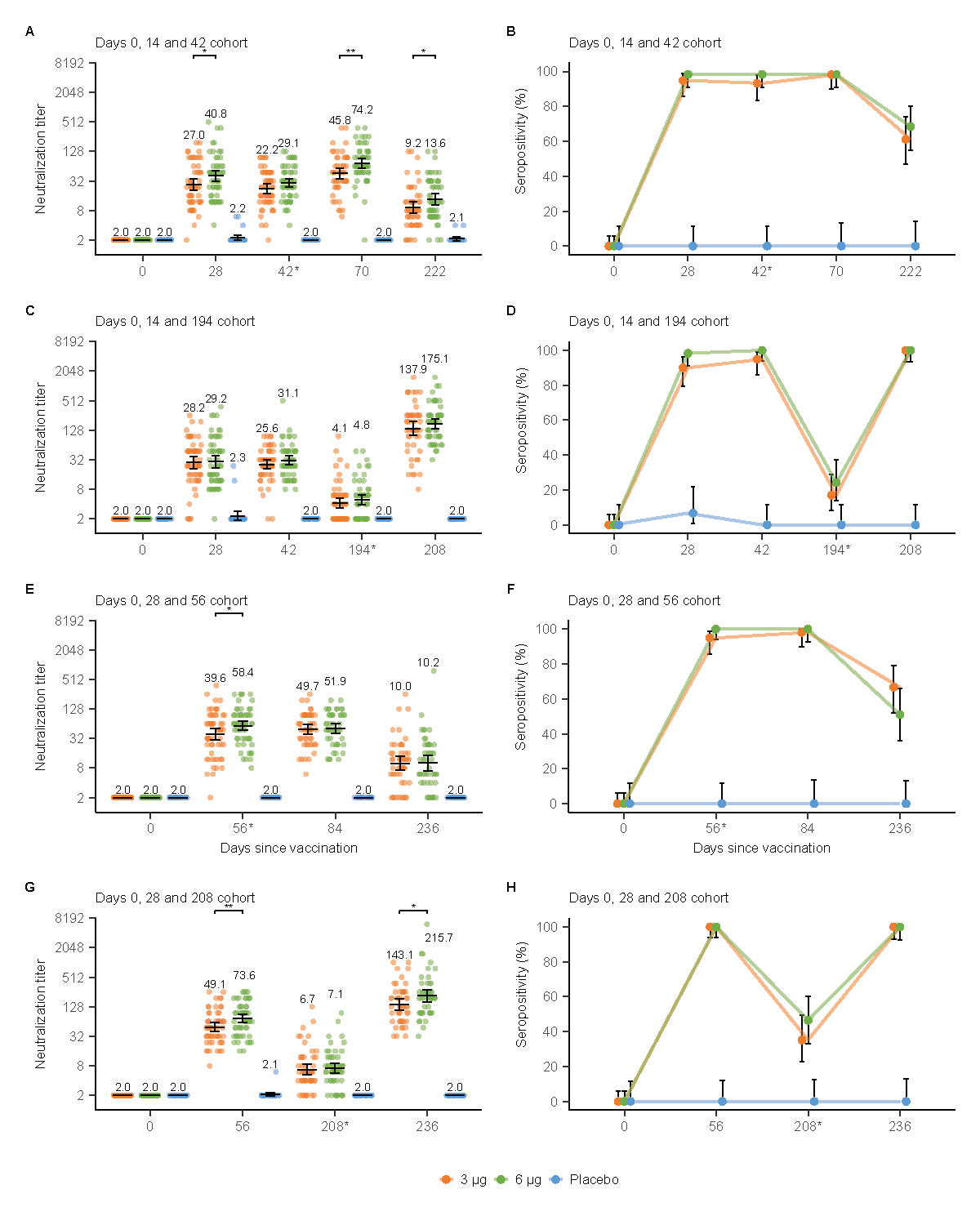

The error bars indicate the 95% CI of the GMT and the spots indicated the individual antibody titres, with the numbers above the spots showing the GMT estimate. Only p values for significant differences are shown on the figure, all p values for all data are in table S1 and table S2. SARS-CoV-2=severe acute respiratory syndrome coronavirus 2.

Supplemental Results of safety

Table S4. Overview of adverse events reported within 28 days post three doses for schedule 1 (n, %)

| **Adverse events** | **3 μg group (N=60)** | **6 μg group (N=60)** | **Placebo**  **(N=30)** | **Total**  **(N=150)** | **P value*** |
| --- | --- | --- | --- | --- | --- |
| Total | 26(43.33) | 25(41.67) | 7(23.33) | 58(38.67) | 0.1559 |
| Vaccine-related† | 22(36.67) | 22(36.67) | 6(20.00) | 50(33.33) | 0.2520 |
| Local | 16(26.67) | 18(30.00) | 3(10.00) | 37(14.67) | 0.0918 |
| Systemic | 11(18.33) | 7(11.67) | 4(13.33) | 22(14.67) | 0.6315 |
| Solicited | 21(35.00) | 21(35.00) | 6(20.00) | 48(32.00) | 0.3142 |
| Unsolicited | 3(5.00) | 1(1.67) | 0(0.00) | 4(2.67) | 0.5309 |
| With 30 minutes | 4(6.67) | 5(8.33) | 0(0.00) | 9(6.00) | 0.3606 |
| Within 7 days | 22(36.67) | 22(36.67) | 6(20.00) | 50(33.33) | 0.2520 |
| Post first dose | 11(18.33) | 11(18.33) | 5(16.67) | 27(18.00) | 1.0000 |
| Post second dose | 9(15.00) | 10(16.67) | 1(3.33) | 20(13.33) | 0.2047 |
| Post third dose | 5(9.09) | 6(10.34) | 0(0.00) | 11(7.91) | 0.2512 |

Note: data in the table were reported within 28 days post any of the three doses (within 14 days post dose 1).

*P value was calculated by Fisher exact probability method and of the comparison of incidence rate among three groups.

†: Vaccine-related adverse events mean that adverse events and vaccines were "possibly related, probably related and definitely related".

Table S5. Adverse reactions reported within 28 days post each dose for schedule 1 (n, %)

| **Adverse Reactions**  **(System organ class, preferred term)** | **Dose 1†** | | | | | **Dose 2** | | | | | **Dose 3** | | | | |
| --- | --- | --- | --- | --- | --- | --- | --- | --- | --- | --- | --- | --- | --- | --- | --- |
|  | **3 μg group (N=60)** | **6 μg group (N=60)** | **Placebo**  **(N=30)** | **Total**  **(N=150)** | **P value*** | **3 μg group (N=60)** | **6 μg group (N=60)** | **Placebo**  **(N=30)** | **Total**  **(N=150)** | **P value*** | **3 μg group (N=55)** | **6 μg group (N=58)** | **Placebo**  **(N=26)** | **Total**  **(N=139)** | **P value*** |
| **Total** | 11(18.33) | 11(18.33) | 5(16.67) | 27(18.00) | 1.0000 | 9(15.00) | 10(16.67) | 1(3.33) | 20(13.33) | 0.2047 | 5(9.09) | 6(10.34) | - | 11(7.91) | 0.2512 |
| Grade 1 | 11(18.33) | 11(18.33) | 5(16.67) | 27(18.00) | 1.0000 | 9(15.00) | 10(16.67) | 1(3.33) | 20(13.33) | 0.2047 | 5(9.09) | 5(8.62) | - | 10(7.19) | 0.3535 |
| Grade 2 | 1(1.67) | - | - | 1(0.67) | 1.0000 | 1(1.67) | 1(1.67) | - | 2(1.33) | 1.0000 | 1(1.82) | 1(1.72) | - | 2(1.44) | 1.0000 |
| General disorders and administration site conditions | 7(11.67) | 10(16.67) | 4(13.33) | 21(14.00) | 0.7544 | 9(15.00) | 9(15.00) | - | 18(12.00) | 0.0478 | 3(5.45) | 5(8.62) | - | 8(5.76) | 0.4014 |
| Injection-site pain | 5(8.33) | 8(13.33) | 3(10.00) | 16(10.67) | 0.7036 | 7(11.67) | 8(13.33) | - | 15(10.00) | 0.0854 | 3(5.45) | 5(8.62) | - | 8(5.76) | 0.4014 |
| Fatigue | 1(1.67) | 1(1.67) | 2(6.67) | 4(2.67) | 0.3282 | 2(3.33) | - | - | 2(1.33) | 0.3557 | - | - | - | - | - |
| Fever | 1(1.67) | 1(1.67) | - | 2(1.33) | 1.0000 | - | - | - | - | - | - | - | - | - | - |
| Injection-site swelling | - | - | - | - | - | - | 2(3.33) | - | 2(1.33) | 0.3557 | - | - | - | - | - |
| Injection-site hypoesthesia | 1(1.67) | 1(1.67) | - | 2(1.33) | 1.0000 | - | - | - | - | - | - | - | - | - | - |
| Injection-site redness | - | - | - | - | - | 1(1.67) | 1(1.67) | - | 2(1.33) | 1.0000 | - | - | - | - | - |
| Injection-site discoloration | - | - | - | - | - | 1(1.67) | - | - | 1(0.67) | 1.0000 | - | - | - | - | - |
| Injection-site itching | - | - | - | - | - | 1(1.67) | - | - | 1(0.67) | 1.0000 | - | - | - | - | - |
| Injection-site induration | - | - | - | - | - | - | 1(1.67) | - | 1(0.67) | 1.0000 | - | - | - | - | - |
| Gastrointestinal disorders | 3(5.00) | - | - | 3(2.00) | 0.2262 | - | - | 1(3.33) | 1(0.67) | 0.2000 | 1(1.82) | 1(1.72) | - | 2(1.44) | 1.0000 |
| Diarrhea | - | - | - | - | - | - | - | - | - | - | 1(1.82) | - | - | 1(0.72) | 0.5827 |
| Nausea | 1(1.67) | - | - | 1(0.67) | 1.0000 | - | - | - | - | - | 1(1.82) | 1(1.72) | - | 2(1.44) | 1.0000 |
| Vomiting | 1(1.67) | - | - | 1(0.67) | 1.0000 | - | - | - | - | - | - | - | - | - | - |
| Nervous system disorders | 1(1.67) | - | 1(3.33) | 2(1.33) | 0.6779 | 1(1.67) | 1(1.67) | - | 2(1.33) | 1.0000 | - | 1(1.72) | - | 1(0.72) | 1.0000 |
| Headache | - | - | 1(3.33) | 1(0.67) | 0.2000 | 1(1.67) | 1(1.67) | - | 2(1.33) | 1.0000 | - | 1(1.72) | - | 1(0.72) | 1.0000 |
| Dizziness | 1(1.67) | - | - | 1(0.67) | 1.0000 | - | - | - | - | - | - | - | - | - | - |
| Musculoskeletal and connective tissue disorders | 2(3.33) | - | - | 2(1.33) | 0.3557 | - | - | - | - | - | 1(1.82) | - | - | 1(0.72) | 0.5827 |
| Muscle pain | 2(3.33) | - | - | 2(1.33) | 0.3557 | - | - | - | - | - | 1(1.82) | - | - | 1(0.72) | 0.5827 |
| Respiratory, thoracic and mediastinal disorders | - | 1(1.67) | - | 1(0.67) | 1.0000 | - | - | - | - | - | - | 1(1.72) | - | 1(0.72) | 1.0000 |
| Cough | - | 1(1.67) | - | 1(0.67) | 1.0000 | - | - | - | - | - | - | 1(1.72) | - | 1(0.72) | 1.0000 |

†: Reported within 14 days post dose 1.

* P value was calculated by Fisher exact probability method and of the comparison of incidence rate among three groups.

Table S6. Overview of adverse events reported within 28 days post the third dose for schedule 2 (n, %)

| **Adverse events** | **3 μg group**  **(N=55)** | **6 μg group**  **(N=56)** | **Placebo**  **(N=30)** | **Total**  **(N=141)** | **P value*** |
| --- | --- | --- | --- | --- | --- |
| Total | 10(18.18) | 13(23.21) | 5(16.67) | 28(19.86) | 0.7637 |
| Vaccine-related† | 10(18.18) | 13(23.21) | 3(10.00) | 26(18.44) | 0.3339 |
| Local | 8(14.55) | 10(17.86) | 2(6.67) | 20(14.18) | 0.4251 |
| Systemic | 3(5.45) | 5(8.93) | 1(3.33) | 9(6.38) | 0.7421 |
| Solicited | 9(16.36) | 13(23.21) | 3(10.00) | 25(17.73) | 0.3076 |
| Unsolicited | 1(1.82) | 3(5.36) | 0(0.00) | 4(2.84) | 0.5359 |
| With 30 minutes | 3(5.45) | 0(0.00) | 1(3.33) | 4(2.84) | 0.2085 |
| Within 7 days | 10(18.18) | 13(23.21) | 3(10.00) | 26(18.44) | 0.3339 |
| 8-28 days | 0(0.00) | 0(0.00) | 0(0.00) | 0(0.00) | 1.0000 |

*P value was calculated by Fisher exact probability method and of the comparison of incidence rate among three groups.

†: Vaccine-related adverse events mean that adverse events and vaccines were "possibly related, probably related and definitely related".

Table S7. Adverse reactions reported within 28 days post the third dose for schedule 2 (n, %)

| **Adverse Reactions**  **(System organ class, preferred term)** | **3 μg group**  **(N=55)** | **6 μg group**  **(N=56)** | **Placebo**  **(N=30)** | **Total**  **(N=141)** | **P value *** |
| --- | --- | --- | --- | --- | --- |
| Total | 10(18.18) | 13(23.21) | 3(10.00) | 26(18.44) | 0.3339 |
| Grade 1 | 10(18.18) | 13(23.21) | 3(10.00) | 26(18.44) | 0.3339 |
| Grade 2 | 1(1.82) | 0(0.00) | 1(3.33) | 2(1.42) | 0.5177 |
| General disorders and administration site conditions | 8(14.55) | 11(19.64) | 2(6.67) | 21(14.89) | 0.2997 |
| Injection-site pain | 8(14.55) | 9(16.07) | 0(0.00) | 17(12.06) | 0.0482 |
| Injection-site itching | 0(0.00) | 1(1.79) | 2(6.67) | 3(2.13) | 0.1145 |
| Fever | 0(0.00) | 1(1.79) | 0(0.00) | 1(0.71) | 1.0000 |
| Fatigue | 0(0.00) | 1(1.79) | 0(0.00) | 1(0.71) | 1.0000 |
| Injection-site swelling | 0(0.00) | 0(0.00) | 1(3.33) | 1(0.71) | 0.2128 |
| Nervous system disorders | 1(1.82) | 2(3.57) | 1(3.33) | 4(2.84) | 1.0000 |
| Headache | 1(1.82) | 2(3.57) | 1(3.33) | 4(2.84) | 1.0000 |
| Dizziness | 0(0.00) | 1(1.79) | 0(0.00) | 1(0.71) | 1.0000 |
| Respiratory, thoracic and mediastinal disorders | 1(1.82) | 2(3.57) | 0(0.00) | 3(2.13) | 0.7979 |
| Cough | 0(0.00) | 2(3.57) | 0(0.00) | 2(1.42) | 0.3506 |
| Laryngeal stimulation | 1(1.82) | 0(0.00) | 0(0.00) | 1(0.71) | 0.6028 |
| Oropharyngeal pain | 0(0.00) | 1(1.79) | 0(0.00) | 1(0.71) | 1.0000 |
| Musculoskeletal and connective tissue disorders | 0(0.00) | 1(1.79) | 0(0.00) | 1(0.71) | 1.0000 |
| Muscle pain | 0(0.00) | 1(1.79) | 0(0.00) | 1(0.71) | 1.0000 |
| Gastrointestinal disorders | 1(1.82) | 0(0.00) | 0(0.00) | 1(0.71) | 0.6028 |
| Nausea | 1(1.82) | 0(0.00) | 0(0.00) | 1(0.71) | 0.6028 |
| Eye disorders | 0(0.00) | 1(1.79) | 0(0.00) | 1(0.71) | 1.0000 |
| Periorbital oedema | 0(0.00) | 1(1.79) | 0(0.00) | 1(0.71) | 1.0000 |

*P value was calculated by Fisher exact probability method and of the comparison of incidence rate among three groups.

Table S8. Overview of adverse events reported within 28 days post three doses for schedule 3 (n, %)

| **Adverse events** | **3 μg group (N=60)** | **6 μg group (N=60)** | **Placebo**  **(N=30)** | **Total**  **(N=150)** | **P value*** |
| --- | --- | --- | --- | --- | --- |
| Total | 20(33.33) | 20(33.33) | 9(30.00) | 49(32.67) | 0.9483 |
| Vaccine-related† | 14(23.33) | 11(18.33) | 8(26.67) | 33(22.00) | 0.6210 |
| Local | 10(16.67) | 7(11.67) | 6(20.00) | 23(15.33) | 0.5336 |
| Systemic | 6(10.00) | 7(11.67) | 3(10.00) | 16(10.67) | 1.0000 |
| Solicited | 14(23.33) | 11(18.33) | 7(23.33) | 32(21.33) | 0.7850 |
| Unsolicited | 0(0.00) | 0(0.00) | 1(3.33) | 1(0.67) | 0.2000 |
| With 30 minutes | 3(5.00) | 2(3.33) | 5(16.67) | 10(6.67) | 0.0529 |
| Within 7 days | 14(23.33) | 11(18.33) | 8(26.67) | 33(22.00) | 0.6210 |
| Post first dose | 11(18.33) | 10(16.67) | 7(23.33) | 28(18.67) | 0.7384 |
| Post second dose | 3(5.08) | 6(10.00) | 2(6.67) | 11(7.38) | 0.6676 |
| Post third dose | 3(5.56) | 1(2.00) | 0(0.00) | 4(3.08) | 0.5305 |

Note: data in the table were reported within 28 days post any of the three doses.

*P value was calculated by Fisher exact probability method and of the comparison of incidence rate among three groups.

†: Vaccine-related adverse events mean that adverse events and vaccines were "possibly related, probably related and definitely related".

Table S9. Adverse reactions reported within 28 days post each dose for schedule 3 (n, %)

| **Adverse Reactions**  **(System organ class, preferred term)** | **Dose 1** | | | | | **Dose 2** | | | | | **Dose 3** | | | | |
| --- | --- | --- | --- | --- | --- | --- | --- | --- | --- | --- | --- | --- | --- | --- | --- |
|  | **3 μg group (N=60)** | **6 μg group**  **(N=60)** | **Placebo**  **(N=30)** | **Total**  **(N=150)** | **P value*** | **3 μg group**  **(N=59)** | **6 μg group**  **(N=60)** | **Placebo**  **(N=30)** | **Total**  **(N=149)** | **P value*** | **3 μg group**  **(N=54)** | **6 μg group**  **(N=50)** | **Placebo**  **(N=26)** | **Total**  **(N=130)** | **P value*** |
| **Total** | 11(18.33) | 10(16.67) | 7(23.33) | 28(18.67) | 0.7384 | 3(5.08) | 6(10.00) | 2(6.67) | 11(7.38) | 0.6676 | 3(5.56) | 1(2.00) | - | 4(3.08) | 0.5305 |
| Grade 1 | 11(18.33) | 9(15.00) | 7(23.33) | 27(18.00) | 0.5763 | 3(5.08) | 6(10.00) | 2(6.67) | 11(7.38) | 0.6676 | 3(5.56) | 1(2.00) | - | 4(3.08) | 0.5305 |
| Grade 2 | - | 2(3.33) | - | 2(1.33) | 0.3557 | - | - | - | - | - | - | - | - | - | - |
| General disorders and administration site conditions | 8(13.33) | 10(16.67) | 5(16.67) | 23(15.33) | 0.8743 | 2(3.39) | 6(10.00) | 2(6.67) | 10(6.71) | 0.3984 | 2(3.7) | 1(2.00) | - | 3(2.31) | 1.0000 |
| Injection-site pain | 7(11.67) | 6(10.00) | 3(10.00) | 16(10.67) | 1.0000 | 2(3.39) | 4(6.67) | 2(6.67) | 8(5.37) | 0.7133 | 1(1.85) | 1(2.00) | - | 2(1.54) | 1.0000 |
| Fatigue | 1(1.67) | 2(3.33) | 1(3.33) | 4(2.67) | 1.0000 | - | 1(1.67) | - | 1(0.67) | 1.0000 | 1(1.85) | - | - | 1(0.77) | 1.0000 |
| Fever | - | 3(5.00) | - | 3(2.00) | 0.2262 | - | 2(3.33) | - | 2(1.34) | 0.5157 | - | - | - | - | - |
| Injection-site discoloration | - | - | 1(3.33) | 1(0.67) | 0.2000 | - | - | - | - | - | - | - | - | - | - |
| Injection-site swelling | - | - | - | - | - | - | - | 1(3.33) | 1(0.67) | 0.2013 | - | - | - | - | - |
| Injection-site redness | - | 1(1.67) | - | 1(0.67) | 1.0000 | - | - | - | - | - | - | - | - | - | - |
| Musculoskeletal and connective tissue disorders | 2(3.33) | 1(1.67) | 1(3.33) | 4(2.67) | 1.0000 | - | - | - | - | - | - | - | - | - | - |
| Muscle pain | 2(3.33) | 1(1.67) | 1(3.33) | 4(2.67) | 1.0000 | - | - | - | - | - | - | - | - | - | - |
| Gastrointestinal disorders | 1(1.67) | - | 1(3.33) | 2(1.33) | 0.6779 | 1(1.69) | 1(1.67) | - | 2(1.34) | 1.0000 | 1(1.85) | - | - | 1(0.77) | 1.0000 |
| Diarrhea | 1(1.67) | - | - | 1(0.67) | 1.0000 | 1(1.69) | - | - | 1(0.67) | 0.5973 | 1(1.85) | - | - | 1(0.77) | 1.0000 |
| Nausea | - | - | - | - | - | - | - | - | - | - | 1(1.85) | - | - | 1(0.77) | 1.0000 |
| Vomiting | - | - | 1(3.33) | 1(0.67) | 0.2000 | - | 1(1.67) | 0(0) | 1(0.67) | 1.0000 | - | - | - | - | - |
| Metabolism and nutrition disorders | - | 1(1.67) | - | 1(0.67) | 1.0000 | - | - | - | - | - | - | - | - | - | - |
| Decreased appetite | - | 1(1.67) | - | 1(0.67) | 1.0000 | - | - | - | - | - | - | - | - | - | - |
| Respiratory, thoracic and mediastinal disorders | - | - | - | - | - | - | - | - | - | - | 2(3.70) | - | - | 2(1.54) | 0.6780 |
| Cough | - | - | - | - | - | - | - | - | - | - | 2(3.70) | - | - | 2(1.54) | 0.6780 |
| Nervous system disorders | - | 1(1.67) | - | 1(0.67) | 1.0000 | - | - | - | - | - | - | - | - | - | - |
| Headache | - | 1(1.67) | - | 1(0.67) | 1.0000 | - | - | - | - | - | - | - | - | - | - |
| Immune system disorders | 1(1.67) | - | - | 1(0.67) | 1.0000 | - | - | - | - | - | - | - | - | - | - |
| Hypersensitivity | 1(1.67) | - | - | 1(0.67) | 1.0000 | - | - | - | - | - | - | - | - | - | - |

*P value was calculated by Fisher exact probability method and of the comparison of incidence rate among three groups.

Table S10. Overview of adverse events reported within 28 days post the third dose for schedule 4 (n, %)

| **Adverse events** | **3 μg group**  **(N=52)** | **6 μg group**  **(N=50)** | **Placebo**  **(N=28)** | **Total**  **(N=130)** | **P value*** |
| --- | --- | --- | --- | --- | --- |
| Total | 10(19.23) | 12(24.00) | 2(7.14) | 24(18.46) | 0.1770 |
| Vaccine-related† | 8(15.38) | 11(22.00) | 2(7.14) | 21(16.15) | 0.2411 |
| Local | 7(13.46) | 7(14.00) | 0(0.00) | 14(10.77) | 0.0844 |
| Systemic | 3(5.77) | 5(10.00) | 2(7.14) | 10(7.69) | 0.8393 |
| Solicited | 8(15.38) | 11(22.00) | 2(7.14) | 21(16.15) | 0.2411 |
| Unsolicited | 1(1.92) | 1(2.00) | 0(0.00) | 2(1.54) | 1.0000 |
| With 30 minutes | 1(1.92) | 3(6.00) | 0(0.00) | 4(3.08) | 0.4393 |
| Within 7 days | 8(15.38) | 11(22.00) | 2(7.14) | 21(16.15) | 0.2411 |
| 8-28 days | 0(0.00) | 0(0.00) | 0(0.00) | 0(0.00) | 1.0000 |

*P value was calculated by Fisher exact probability method and of the comparison of incidence rate among three groups.

†: Vaccine-related adverse events mean that adverse events and vaccines were "possibly related, probably related and definitely related".

Table S11. Adverse reactions reported within 28 days post the third dose for schedule 4 (n, %)

| **Adverse Reactions**  **(System organ class, preferred term)** | **3 μg group**  **(N=52)** | **6 μg group**  **(N=50)** | **Placebo**  **(N=28)** | **Total**  **(N=130)** | **P value*** |
| --- | --- | --- | --- | --- | --- |
| Total | 8(15.38) | 11(22.00) | 2(7.14) | 21(16.15) | 0.2411 |
| Grade 1 | 7(13.46) | 10(20.00) | 1(3.57) | 18(13.85) | 0.1317 |
| Grade 2 | 1(1.92) | 1(2.00) | 1(3.57) | 3(2.31) | 1.0000 |
| General disorders and administration site conditions | 8(15.38) | 9(18.00) | 1(3.57) | 18(13.85) | 0.1886 |
| Injection-site pain | 6(11.54) | 7(14.00) | 0(0.00) | 13(10.00) | 0.1074 |
| Fever | 1(1.92) | 1(2.00) | 1(3.57) | 3(2.31) | 1.0000 |
| Fatigue | 1(1.92) | 2(4.00) | 0(0.00) | 3(2.31) | 0.6112 |
| Injection-site swelling | 1(1.92) | 0(0.00) | 0(0.00) | 1(0.77) | 1.0000 |
| Injection-site itching | 1(1.92) | 0(0.00) | 0(0.00) | 1(0.77) | 1.0000 |
| Nervous system disorders | 1(1.92) | 1(2.00) | 1(3.57) | 3(2.31) | 1.0000 |
| Headache | 1(1.92) | 1(2.00) | 1(3.57) | 3(2.31) | 1.0000 |
| Gastrointestinal disorders | 1(1.92) | 2(4.00) | 0(0.00) | 3(2.31) | 0.6112 |
| Nausea | 0(0.00) | 2(4.00) | 0(0.00) | 2(1.54) | 0.1912 |
| Diarrhea | 1(1.92) | 0(0.00) | 0(0.00) | 1(0.77) | 1.0000 |
| Respiratory, thoracic and mediastinal disorders | 1(1.92) | 1(2.00) | 0(0.00) | 2(1.54) | 1.0000 |
| Oropharyngeal pain | 1(1.92) | 0(0.00) | 0(0.00) | 1(0.77) | 1.0000 |
| Running nose | 0(0.00) | 1(2.00) | 0(0.00) | 1(0.77) | 0.6000 |
| Musculoskeletal and connective tissue disorders | 0(0.00) | 1(2.00) | 0(0.00) | 1(0.77) | 0.6000 |
| Muscle pain | 0(0.00) | 1(2.00) | 0(0.00) | 1(0.77) | 0.6000 |

*P value was calculated by Fisher exact probability method and of the comparison of incidence rate among three groups.

Table S12. Serious adverse events reported (n, %)

| **Adverse events (MedDRA 23.0)** | **3 μg group** | **6 μg group** | **Placebo** | **Total** | **P value^*^** |
| --- | --- | --- | --- | --- | --- |
| ***Schedule 1*** | **N=60** | **N=60** | **N=30** | **N=150** |  |
| Total | 1(1.67) | 2(3.33) | 0(0.00) | 3(2.00) | 0.8041 |
| Hepatobiliary disorders | 1(1.67) | 0(0.00) | 0(0.00) | 1(0.67) | 1.0000 |
| Autoimmune hepatitis | 1(1.67) | 0(0.00) | 0(0.00) | 1(0.67) | 1.0000 |
| Injury, poisoning and procedural complications | 0(0.00) | 1(1.67) | 0(0.00) | 1(0.67) | 1.0000 |
| Ankle fractures | 0(0.00) | 1(1.67) | 0(0.00) | 1(0.67) | 1.0000 |
| Neoplasms benign, malignant and unspecified (incl cysts and  polyps) | 0(0.00) | 1(1.67) | 0(0.00) | 1(0.67) | 1.0000 |
| Teratoma | 0(0.00) | 1(1.67) | 0(0.00) | 1(0.67) | 1.0000 |
| Uterine leiomyoma | 0(0.00) | 1(1.67) | 0(0.00) | 1(0.67) | 1.0000 |
| Reproductive system and breast disorders | 0(0.00) | 1(1.67) | 0(0.00) | 1(0.67) | 1.0000 |
| Pelvic adhesions | 0(0.00) | 1(1.67) | 0(0.00) | 1(0.67) | 1.0000 |
| Gastrointestinal disorders | 1(1.67) | 0(0.00) | 0(0.00) | 1(0.67) | 1.0000 |
| Erosive gastritis | 1(1.67) | 0(0.00) | 0(0.00) | 1(0.67) | 1.0000 |
| Gastroesophageal reflux disease | 1(1.67) | 0(0.00) | 0(0.00) | 1(0.67) | 1.0000 |
| ***Schedule 2*** | **N=60** | **N=60** | **N=30** | **N=150** |  |
| Total | 2(3.33) | 2(3.33) | 0(0.00) | 4(2.67) | 0.6855 |
| Injury, poisoning and procedural complications | 1(1.67) | 1(1.67) | 0(0.00) | 2(1.33) | 1.0000 |
| Soft tissue injury | 0(0.00) | 1(1.67) | 0(0.00) | 1(0.67) | 1.0000 |
| Hand fracture | 1(1.67) | 0(0.00) | 0(0.00) | 1(0.67) | 1.0000 |
| Nail injury | 1(1.67) | 0(0.00) | 0(0.00) | 1(0.67) | 1.0000 |
| Renal and urinary disorders | 0(0.00) | 1(1.67) | 0(0.00) | 1(0.67) | 1.0000 |
| Cystitis glandularis | 0(0.00) | 1(1.67) | 0(0.00) | 1(0.67) | 1.0000 |
| Gastrointestinal disorders | 1(1.67) | 0(0.00) | 0(0.00) | 1(0.67) | 1.0000 |
| Hemorrhoid | 1(1.67) | 0(0.00) | 0(0.00) | 1(0.67) | 1.0000 |
| ***Schedule 3*** | **N=60** | **N=60** | **N=30** | **N=150** |  |
| Total | 0(0.00) | 1(1.67) | 0(0.00) | 1(0.67) | 1.0000 |
| Surgical and medical procedures | 0(0.00) | 1(1.67) | 0(0.00) | 1(0.67) | 1.0000 |
| Induced abortion | 0(0.00) | 1(1.67) | 0(0.00) | 1(0.67) | 1.0000 |
| ***Schedule 4*** | **N=60** | **N=60** | **N=30** | **N=150** |  |
| Total | 1(1.67) | 0(0.00) | 0(0.00) | 1(0.67) | 1.0000 |
| Musculoskeletal and connective tissue disorders | 1(1.67) | 0(0.00) | 0(0.00) | 1(0.67) | 1.0000 |
| Herniated intervertebral disc | 1(1.67) | 0(0.00) | 0(0.00) | 1(0.67) | 1.0000 |

*P value was calculated by Fisher exact probability method and of the comparison of incidence rate among three groups.
